# Functionality and phenotype of T cells in patients with varying severity of acute dengue and metabolic status

**DOI:** 10.1101/2025.07.02.25330735

**Authors:** Heshan Kuruppu, Malithi De Silva, Chathumini Dissanayake, Radandee Rathnapriya, Ananda Wijewickrama, Damayanthi Idampitiya, Chandima Jeewandara, Graham Ogg, Gathsaurie Neelika Malavige

## Abstract

**Background:** Currently the role of dengue virus (DENV) specific T cell responses in disease pathogenesis and protection are not well understood, including potential differences in those who have obesity. We sought to investigate the functionality and phenotype of T cells in patients with acute dengue fever (DF) or dengue haemorrhagic fever (DHF).

**Methods:** T cell function was assessed in patients with DF (n=50) and DHF (n=12), recruited within ≤4 days of illness and again on day 5 to 7, using 109 peptides representing CD8□ epitopes and 90 peptides targeting CD4+ T cell epitopes. Phenotypic analysis was in DF (n=21) and DHF patients (n=21), recruited between days 6–8 since onset of illness, by multicolor flow cytometry.

**Results:** The frequency of ex vivo IFNγ ELISpot responses to both the CD4+ and CD8+ peptides pools significantly increased from the first to second time point in patients with DF (p<0.0001) but not with DHF. The frequency of ex vivo IFNγ ELISpot responses to CD4+ (p=0.001) and CD8+ peptides pools (p=0.0002) also significantly increased from the first to second time point in lean patients compared obese patients. Cutaneous lymphocyte associated antigen (CLA) expression was significantly higher in the CD8+ T cell subset in patients with DF and DHF compared to HC and these differences were most significant in CD8+CD45RA− T cells. CD8+CD45RA-CLA+ T cells consisted of >50% of the T cells in 9/21 patients with DHF, with 92.7% expressing CD38. CLA expression was highest in the CD8+CD45RA− of obese individuals, which was significantly higher compared to lean individuals (p=0.01). CD27 and CD127 were both significantly downregulated in patients with DHF compared to DF, with ICOS expression being significantly higher in CD8+ T cells in DHF.

**Discussion:** Patients with DHF and obese individuals had impaired T cell functionality. Activated and skin homing CD8+ T cells were associated with DHF, with downregulation of CD27 and CD127. Therefore, the role of skin homing T cells, which have impaired functionality in disease pathogenesis, should be further investigated.

## Introduction

Dengue infections are one of the most rapidly emerging mosquito-borne viral infections, with 5.66 billion of the world population estimated to be at risk of infection with the dengue virus (DENV) [21]. Due to high transmission rates in recent years causing large outbreaks in many countries, the DENV has been named a pathogen with a pandemic potential in World Health Organization R&D blueprint published in 2024 [34]. While most individuals with dengue infection experience an asymptomatic or mild illness, some individuals develop plasma leakage, bleeding and organ dysfunction, resulting in dengue haemorrhagic fever (DHF). Although the incidence of dengue has been rising in all age groups, symptomatic infection has particularly increased in individuals aged 15 to 49 years in all countries [43]. However, the mortality rates have been rising in individuals aged 50 years and above, especially in countries which belong to the high and middle socio-demographic index [43]. Older age, along with the presence of comorbidities such as diabetes, renal disease and hypertension have shown to increase the risk of severe disease, which possibly attributes to higher mortality rates in older individuals [31].

Currently the reasons why some individuals develop mild or asymptomatic illness while others develop plasma leakage, organ dysfunction and bleeding when infected with the DENV are not entirely understood. Many factors such as genetic susceptibility, presence of comorbidities leading to a dysfunctional immune response, prior exposure to flaviviruses and the magnitude and the functionality of antibody and T cell responses are likely to contribute to disease pathogenesis [10,26,27,38]. While early appearance of DENV NS3 and NS5 specific polyfunctional T cell responses [40,41] and CD4+ and CD8+ T cells restricted through specific HLAs [36,37] have shown to associate with milder forms of dengue, with cross reactive T cell responses [8], CD4+ T cells lack CXCR5, with high expression of PD-1 [2], effector memory T cells re-expressing CD45RA (Temra) have shown to associate with severe disease [30]. Furthermore, a higher magnitude of T cell responses following DENV infection, has been shown to protect against symptomatic infection during subsequent infection [10,24]. Given that recent vaccine trials show that the presence of neutralizing antibodies alone do not confer complete protection against dengue [27], it is becoming increasingly important to understand T cell responses that associate with protection vs disease pathogenesis.

Obesity is a risk factor for severe disease in COVID-19, influenza and many other viral infections, attributed to a dysfunctional adipose tissue and adipokine profile, altered monocyte and mast cell phenotypes, increase in mast cells, chronic low-grade inflammation, dysfunctional NK and T-cell responses and waning of neutralizing antibodies [1,7,35]. Obese mice have shown to have higher viral loads and an impaired CD8+ T cell response, with reduced IFNγ production during influenza infection [16]. Furthermore, obese individuals had lower frequencies of influenza specific CD8+ T cell responses with impaired IFNγ and granzyme B production following influenza vaccination [32]. We recently showed that obese individuals were more likely to have DHF, elevated liver enzymes and higher levels of C-reactive protein and ferritin than lean individuals [19]. As obesity and associated metabolic diseases such as diabetes and cardiovascular disease are risk factors for severe dengue [31], it would be important to investigate the differences in the functionality and phenotype of T cell responses associated with clinical disease severity and in obesity to better understand disease pathogenesis. Therefore, in this study we assessed the functionality of DENV-specific T cell responses using peptides specific to 109 CD8+ and 90 CD4+ T cell epitopes identified from all four DENV serotypes, in patients with varying severity of disease severity and obesity. We further assessed the phenotypes of CD4+ and CD8+ T cell subsets in patients with DF and DHF, in lean and obese individuals comparing them with healthy controls to understand the T cell responses that are associated with clinical disease severity and obesity.

## Methods

### Patient recruitment and classification of disease severity

A total of 104 adult patients with acute dengue infection were recruited from the National Institute of Infectious Diseases, Sri Lanka, between May 2023 and March 2025 following informed written consent. Patients with chronic kidney or liver disease were excluded. Clinical characteristics, laboratory parameters, and ultrasound scans were recorded daily and used to assess the presence of fluid leakage and disease severity. Patients were classified as having DF or DHF according to WHO 2011 criteria [39].

Due to the limitations in the blood volume that could be obtained from a patient (as they were bled multiple times a day as a part of their routine management), we initially carried out *ex vivo* IFNγ ELISpot assays in (n=62) patients (cohort 1). In cohort 1, the first blood sample (F) was obtained from ≤4 days from onset of illness and a second blood sample (S) 2 days later (5 to 7 days since onset of illness). None of patients had plasma leakage at the time of recruitment, and were classified as having DHF, as they developed plasma leakage during subsequent follow-up.

Phenotypic analysis of T cells by flowcytometry was carried out in the 2^nd^ cohort (n=42), who were recruited from 6 to 8 days from onset of illness. All patients in cohort 1 had either a positive NS1 antigen test (SD Biosensor, South Korea) or gave a positive result by DENV-specific real-time PCR. In cohort 2, as patients were recruited at a later time point in illness, many patients were not positive for either the dengue NS1 antigen or PCR and therefore, those who had clinical features of dengue, with a positive dengue specific IgM and IgG antibodies were recruited (Abbott Bioline, South Korea).

### Healthy individuals

Healthy adult volunteers were recruited from staff of the University of Sri Jayewardenepura, following informed written consent.

### Ethics statement

This study was approved by the Ethics Review Committee of the Faculty of Medical Sciences, University of Sri Jayewardenepura, Sri Lanka (Approval No. 58/19). Informed written consent was obtained from all participants.

### Anthropometric measurements

At recruitment, the weight was measured using a digital scale, while wearing light clothing and barefooted. The height was measured barefooted, using a stadiometer. Waist circumference was recorded using a non-stretchable measuring tape. Overweight was defined as BMI ≥23.9 kg/m² [5], and central obesity as waist circumference ≥80 cm in women or ≥90 cm in men [33].

### Realtime PCR for detection of the DENV

Viral RNA was extracted from patient serum using the QIAamp Viral RNA Mini Kit (Qiagen, USA). Multiplex real-time PCR was performed using CDC primers and dual-labeled probes to identify DENV serotypes 1–4 (Life Technologies, India) as described previously [3].

### Peptide pools used in the ex vivo IFN**γ** ELISpot assays

Peripheral blood mononuclear cells (PBMCs) were separated from whole blood using density gradient centrifugation with Lymphoprep™. Cells were stimulated with a peptide pool, which contained previously described immunodominant CD4+ and CD8+ T cell epitopes (Mabtech, Cat: 3698-1 and 3695-1). This pool of peptides included 109 peptides representing CD8 epitopes (8 to 9aa long) across the whole DENV polypeptide: Capsid (9 peptides), Envelope (9 peptides), NS1 (3 peptides), NS2A (5 peptides), NS2B (6 peptides), NS3 (41 peptides), NS4A (5 peptides), NS4B (9 peptides), and NS5 (22 peptides). The CD8+ T cell epitopes were shown to be restricted through HLA class I A1, A2, A3, A24, B7, B44, and B8 alleles, identified from DENV-1 (31 peptides), DENV-2 (55 peptides), DENV-3 (19 peptides), and DENV-4 (15 peptides). The pool also contained 90 peptides targeting CD4+ T cell epitopes specific to capsid (37 peptides), membrane (2 peptides), envelope (11 peptides), NS1 (3 peptides), NS2A (3 peptides), NS2B (4 peptides), NS3 (25 peptides), NS4A (1 peptide), NS4B (2 peptides), and NS5 (2 peptides). The CDC+ T cell epitopes were shown to be restricted through HLA class II alleles DRB1, DRB5, DRB3, DRB4, DQA1, DQB1, and DPB1, identified from DENV-1 (14 peptides), DENV-2 (60 peptides), DENV-3 (15 peptides), and DENV-4 (11 peptides).

### Ex vivo IFN**γ** ELISpot assays

Ex vivo IFNγ ELISpot assays were carried out using freshly isolated peripheral blood mononuclear cells (PBMCs) as previously described [25]. 100,000 PBMCs/well were added with the two pools of peptides (CD4+ and CD8+ T cell epitope pools) at a final concentration of 10 µM and incubated overnight. PHA was included as positive control and media alone was used as a negative control. After 16 hours of incubation, IFN-γ-producing cells were detected using a human IFN-γ ELISpot kit (Mabtech, Cat: 3420-2APW-2). The spots were enumerated using an automated ELISpot reader (AID Germany). Background (PBMCs plus media alone) was subtracted and data expressed as number of spot-forming units (SFU) per 10^6^ PBMCs. All experiments were done in duplicate.

### Phenotypic analysis of CD4+ and CD8+ T cells using flow cytometry

To assess the differences in T cell phenotypes in relation to disease severity and obesity, two multicolor flow cytometry panels were used to stain whole blood samples in patients from cohort 2. Panel 1 included markers related to memory and homing (CD45RA, CCR7, CD62L) (BioLegend, USA) and activation (CD38) (BioLegend, USA), along with other markers (CD3, CD4, CD8, CD45) (BioLegend, USA). Panel 2 included additional functional and activation markers such as CD127, CD27 and ICOS (BioLegend, USA). Antibody titrations were performed prior to staining to determine optimal concentrations. After surface staining and red blood cell were lysed using lysing solution (BD FACS lysing solution) and cells were acquired using a BD FACSAria III cytometer (BD Biosciences). FlowJo software was used for data analysis, and the T cell phenotypes were compared across clinical severity groups and obesity status. The gating strategies used for identification of the different markers are shown in supplementary figure S1-S2.

### Quantification of adiponectin levels

Serum adiponectin was measured in patients from both cohorts and healthy individuals using a commercial ELISA kit (Abcam, UK). Samples were stored at −80°C and diluted according to the manufacturer’s instructions. Concentrations were calculated using a four-parameter logistic (4PL) curve.

### Quantification of dengue NS1 antigen levels

Serum NS1 levels were measured in dengue patients using a commercial ELISA kit (ARG81357, Arigo Biolaboratories, Taiwan), following the manufacturer’s instructions. Samples were stored at –80 °C and diluted with 1X PBS when necessary. Standards were prepared by serial dilution to generate a 7-point standard curve ranging from 1.5625 to 100 ng/mL. Absorbance was read at 450 nm, and concentrations were calculated using a four-parameter logistic (4PL) curve.

### Statistical analysis and post-hoc analysis / downstream bioinformatics analysis

Statistical analyses were conducted using GraphPad Prism version 10.4.1 (Dotmatics, California, USA). As the data were not normally distributed, results are presented as median with interquartile range (IQR). For comparisons between two unpaired groups, the two-tailed Mann– Whitney U test was used. For comparisons involving more than two groups, the Kruskal–Wallis test followed by Dunn’s multiple comparison test was applied.

For the downstream bioinformatics analysis, a dataset was constructed comprising 53 samples and 49 flow cytometry markers representing various T-cell subsets and activation states. Disease severity (categorized as Healthy, Dengue Fever [DF], and Dengue Hemorrhagic Fever [DHF]) was used as the primary grouping variable for the analysis.

Data preprocessing included standardization of continuous variables and multicollinearity assessment using variance inflation factors (VIF) and correlation analysis [18]. Marker selection combined statistical criteria (VIF thresholds and correlation patterns) with biological relevance, resulting in 33 flow cytometry markers for analysis.

Unsupervised analyses were conducted to explore patterns in T-cell phenotypes across patient groups. Principal component analysis (PCA) was performed on 33 flow cytometry markers to reduce dimensionality and provide a global overview of immune cell variation among samples [23]. Clustering analyses were then carried out using both K-means and hierarchical approaches, focusing on three biologically defined marker subsets representing distinct functional T-cell states relevant to dengue pathogenesis: skin-homing memory, co-stimulatory, and activation-associated profiles [6,11]. Further details on data preprocessing, dimensionality reduction, and clustering validation are provided in the Supplementary Methods.

## Results

### *Ex vivo* ELISpot responses in patients with varying severity of acute dengue

*Ex vivo* ELISpot assays were carried out in patient cohort 1 (≤ 4 days since onset of symptoms), which consisted of patients with DF (n=50), DHF (n=12) and healthy controls (n=19), supplementary table 1. The DENV was detected in 26/62 (41.9%) samples, with DENV2 in 7 samples and DENV3 in 19 samples, as DENV3 was the predominant circulating serotype during this time [3]. The CD8+ peptide pool had T cell epitopes derived from DENV-1 (31 peptides), DENV-2 (55 peptides), DENV-3 (19 peptides), and DENV-4 (15 peptides) and the CD4+ peptide pool had T cell epitopes derived from DENV-1 (14 peptides), DENV-2 (60 peptides), DENV-3 (15 peptides), and DENV-4 (11 peptides). Therefore, the peptide pools represented all four DENV serotypes. The frequency of *ex vivo* IFNγ ELISpot responses to both the CD4+ and CD8+ peptides pools significantly increased from the first (F) to second (S) time point in patients with DF but not with DHF (Figure 1A). Interestingly, the frequency of IFNγ ELISpot responses to the pool of CD8+ T cell epitopes were significantly higher in early illness in patients with DHF compared to DF (p=0.04).

**Figure 1:**
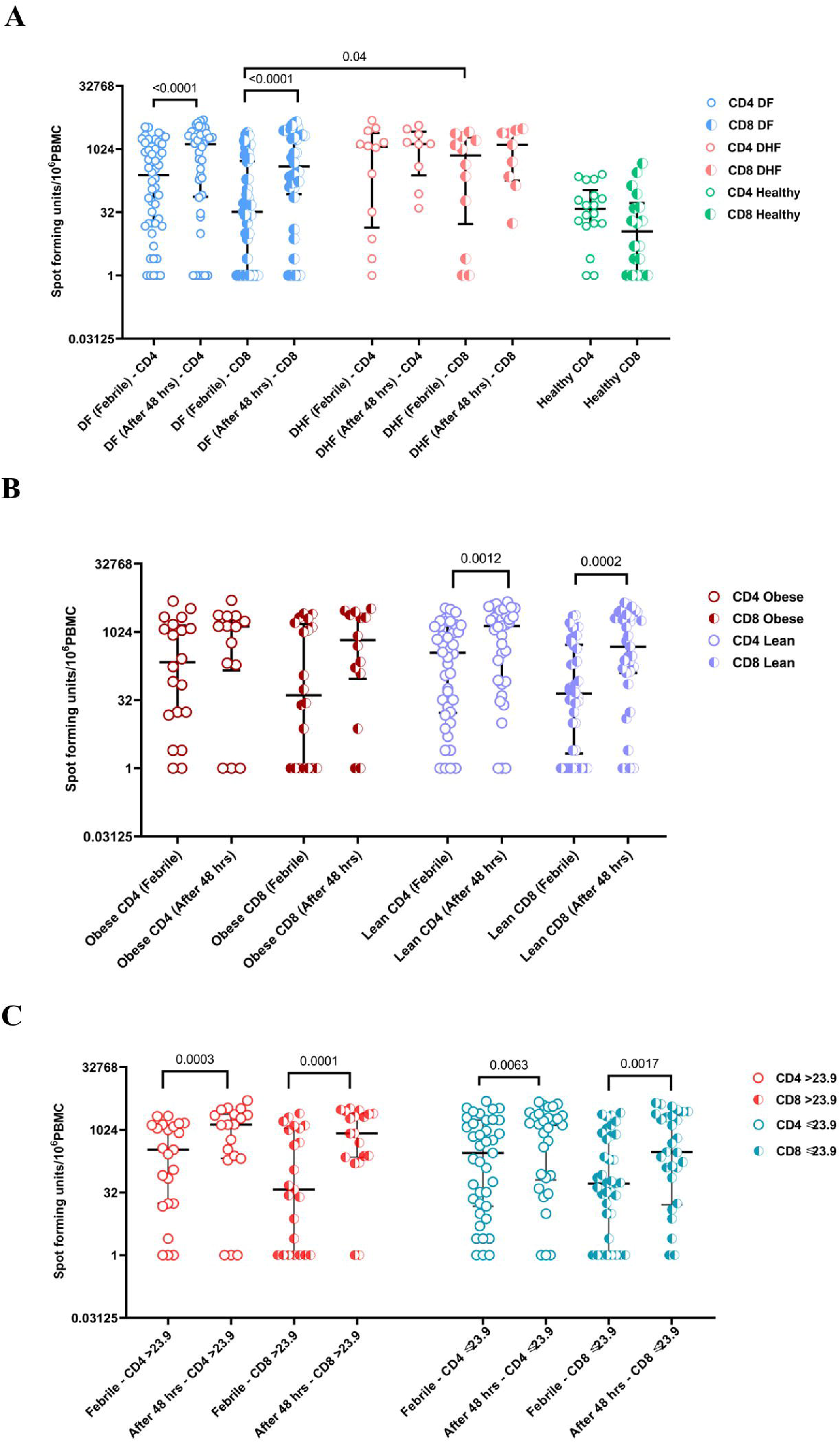
The frequency of CD4+ and CD8+ DENV specific IFN**γ** producing T cells in patients with varying severity of acute dengue and metabolic status. The frequency of DENV-specific CD4+ and CD8+ T cell responses were measured by ex vivo IFNγ ELISpot assays, by using a pool of peptides representing CD8+ T cells epitopes and CD4+ T cell epitopes, in patients with DF (n=50), DHF (n=12) and healthy controls (HC=18) during two time points (F: ≤ day 4 since onset and S: 5-7 days since onset of illness) (A). The frequency of DENV-specific CD4+ and CD8+ T cell responses were also compared in obese (n= 20), vs lean (n= 42) patients (B) and in those with high BMI (>23.9, n= 23) or normal BMI (≤23.9, n= 39) (C) at time point F and S. The Wilcoxon matched pairs signed rank test was used to compare differences in responses from time point F and S, while the Mann Whitney U test (two tailed) was used for comparisons between groups. Error bars indicate the median and interquartile ranges.

As we recently showed that obese individuals had significantly higher levels of inflammatory markers and liver damage, we sought to investigate the differences in T cell functionality in those who were obese compared to lean individuals. We found that the frequency of ex vivo IFNγ ELISpot responses to both the CD4+ and CD8+ peptides pools significantly increased from the first to second time point in lean patients compared obese patients (waist circumference ≥80 cm in women or ≥90 cm in men), (Figure 1B). We also analyzed our data based on the BMI of patients, and all patients (BMI ≤23.9 and BMI >23.9), had a significant increase in the frequency of ex vivo IFNγ ELISpot responses to both the CD4+ (p=0.001) and CD8+ peptides pools (p=0.0002) between the first and second time point of illness (Figure 1C).

### Phenotypic differences in CD4+ and CD8+ T cells in patients with varying severity of acute dengue

Phenotypic analysis of CD4+ and CD8+ T cell subsets were carried out in patient cohort 2 (recruited between day 6 to 8 days since onset of symptoms), which consisted of patients with DF (n=21), DHF (n=21) and healthy controls (n=11), supplementary table 2. There were no differences in proportion of CD4+ or CD8+ T cells, or CD4+ and CD8+ T cells expressing CD45RA+ in patients with DF and DHF compared to healthy individuals (Figure 2A). There was no difference in proportion of CD4+ and CD8+ central memory (CD45RA^−^, CCR7+CD62L+) T cells in healthy controls (HCs), patients with DF and DHF, and also no differences in the proportion of CD4+ effector memory T cells (CD45RA-CCR7-CD62L-) in HCs and patients with DF or DHF. However, patients with DHF had significantly lower portions (p=0.004) of CD8+ effector memory T cells (CD45RA-CCR7-CD62L-), compared to HCs (Figure 2B).

**Figure 2:**
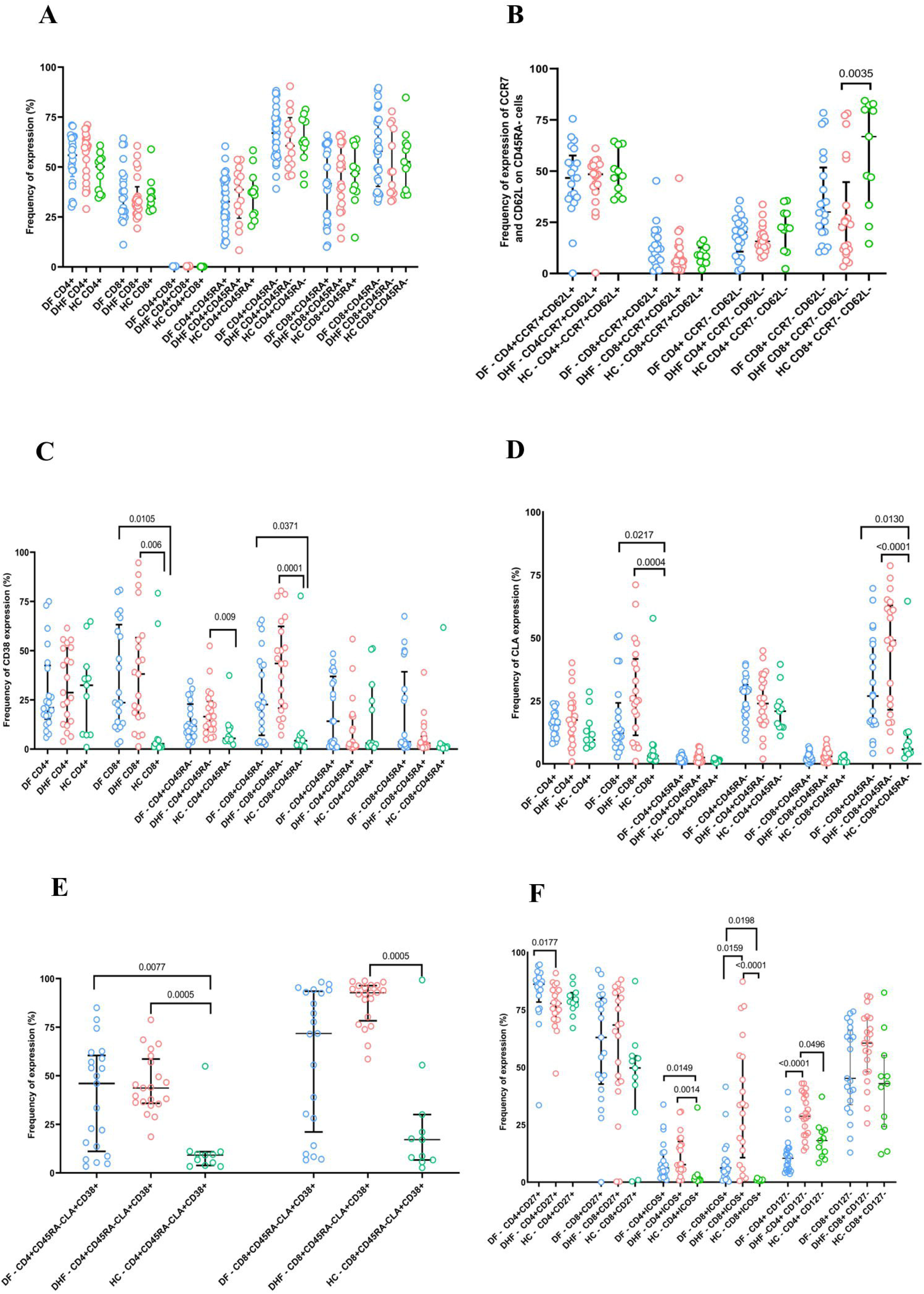
Differences in the phenotype of CD4+ and CD8+ T cells in patients with varying severity of acute dengue. CD4+ and CD8+ T cell subsets were assessed in patients with DF (n=21), DHF (n=21) and healthy controls (HC=11) recruited between days 6–8 from symptom onset by multicolour flowcytometry. The differences between the proportion of CD4+ CD8+ and CD45RA expressing T cells (A), expression of CCR7 and CD62L on CD45RA− T cells (B), CD38 expression (C), CLA expression (D), CD38 expression on CD45RA-CLA+ T cells (E) and CD27, CD127 and ICOS expression were assessed (F). The Kruskal Wallis test followed by Dunn’s multiple comparison test was used to calculate the differences between these cohorts. The error bars indicate the median and the interquartile ranges.

CD38 expression was significantly higher in CD8+ T cells and specifically CD8+CD45RA− T cells in patients with DF and DHF compared to HCs (Figure 2C). For CD4+ T cells, the difference in CD38+ expression was only seen between patients with DHF and HC in CD8+CD45RA− T cells. CLA expression was also significantly higher in the CD8+ T cell subset in patients with DF and DHF compared to HC and these differences were most significant in CD8+CD45RA− T cells (Figure 2D). Although CLA was higher in CD8+CD45RA− T cells of patients with DHF compared to DF, this was not significant. Interestingly, CD8+CD45RA-CLA+ T cells consisted of >50% of the T cells in 9/21 patients with DHF, while a further 4 patients had expression levels between 45-50%. 92.7% of CD8+CD45RA-CLA+ cells also expressed CD38 (IQR 78.3 to 96.4% of cells), which was significantly higher than expression seen in HCs (median 17.1%, IQR 6.4% to 30% of cells) (Figure 2E). 46% of CD4+CD45RA-CLA+ T cells in patients with DF (IQR 11.1 to 60.45% of cells), and 43.7% of CD4+CD45RA-CLA+ T cells of patients with DHF (IQR 35.8% to 58.6% of cells) also expressed CD38, although the expression levels were lower than in CD8+ T cells.

Significant differences in CD27 expression were seen in CD4+ T cells, with CD27+ CD4+ T cells significantly downregulated in CD4+ T cells in patients with DHF compared to DF (Figure 2F). ICOS expression was significantly upregulated in both CD4+ and CD8+ T cells in patients compared to HCs, with CD8+ T cells in patients with DHF having significantly more ICOS expression than patients with DF (Figure 2F). Downregulation of CD127 (IL-7Rα) was higher in both CD8+ and CD4+ T cells in patients with DHF compared to patients with DF and HCs, although these differences were only significant for CD4+ T cells (Figure 2F). We did not observe significant expression of either CD32 or CD64 in either CD4+ T cells or CD8+ T cells in the patients or in HCs (supplementary figure S3).

### Principal component analysis of T cell phenotypic markers that associate with disease severity

Principal component analysis (PCA) of the 33 flow cytometry markers showed that the first two principal components captured 26.31% and 12.55% of the total variance, respectively (Figure 3A). The PCA scatter plot revealed substantial overlap between the three clinical groups (DF, DHF and HCs), though some grouping patterns were seen. Healthy controls (green) showed a tendency to cluster together, particularly in the left quadrants of the plot, suggesting some immunological distinction from dengue-infected patients. However, DF (red) and DHF (orange) samples showed extensive overlap throughout the reduced-dimensional space with no clear separation between disease severity categories. The overall distribution indicated that while the selected immunological markers could partially distinguish healthy individuals from dengue patients, they did not effectively discriminate between different severities of dengue infection in an unsupervised manner.

**Figure 3:**
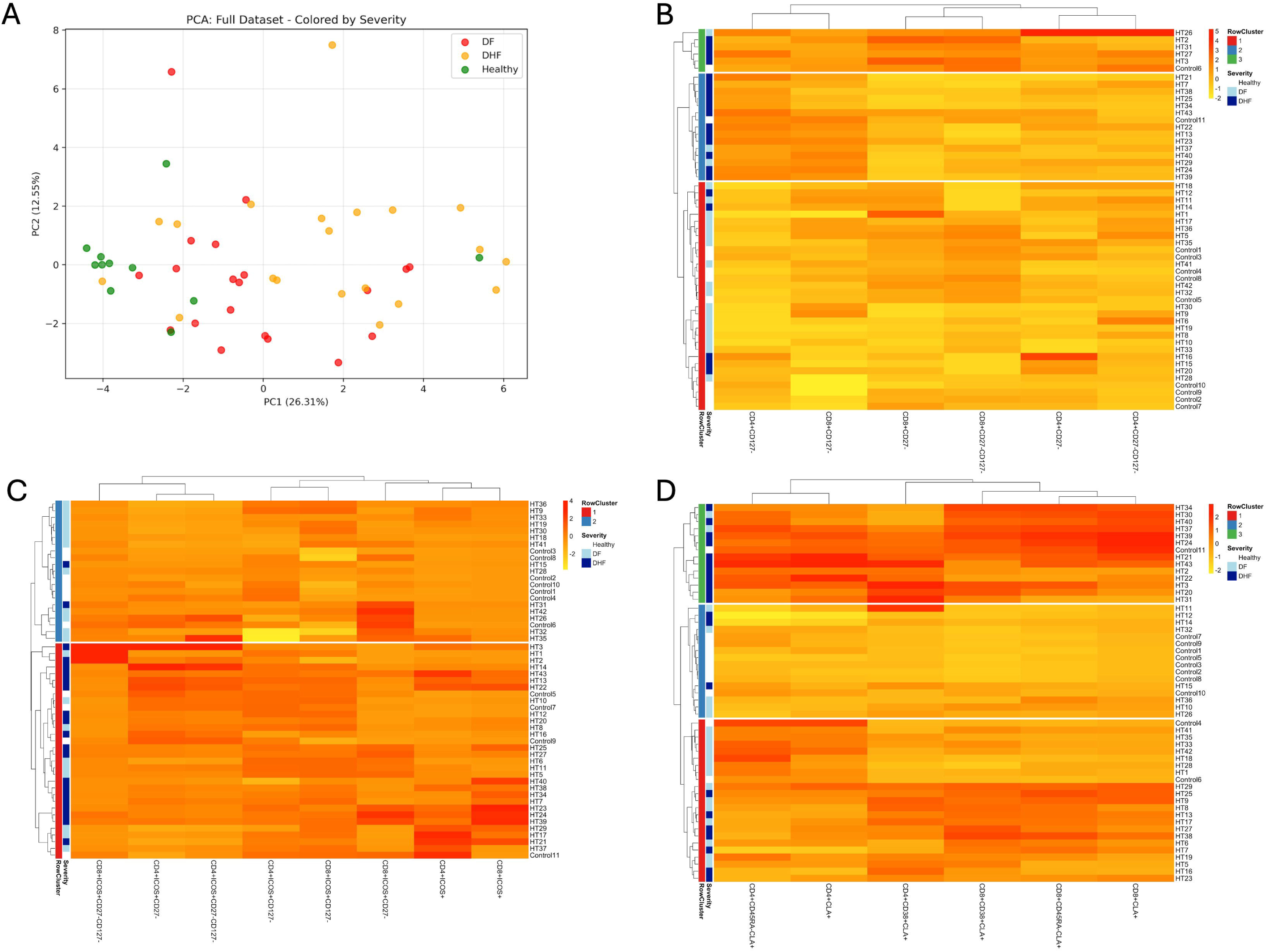
Unsupervised analysis of T cell phenotypic markers by dengue disease severity. The PCA of 33 immunological markers with disease severity is shown. Each point represents an individual sample colored by clinical group: healthy controls (green), DF (red), and DHF (orange). The first two principal components are shown, with PC1 and PC2 explaining 26.31% and 12.55% of the total variance, respectively (A). Hierarchical clustering heatmaps of phenotypic markers associated with co-stimulation and survival (B), expression of ICOS and CD27 (C), CLA (D) is shown. Clustering was performed using Ward’s, average or complete linkage, with color annotations indicating clinical severity and unsupervised cluster assignment. Columns represent marker combinations; rows represent individual samples.

### Hierarchical clustering heatmaps of T cell phenotypic markers that are associated with disease severity

To better understand immune phenotypic patterns, we performed clustering analyses on three biologically relevant T-cell marker subsets, each representing functionally distinct immune states involved in dengue pathogenesis. The optimal clustering configuration for each analysis was determined primarily based on the highest ARI, with silhouette scores considered secondarily for interpretability.

### Phenotypic markers associated co-stimulation and survival

Analysis of the six markers (CD27 and CD127 combinations) using hierarchical clustering with Ward’s linkage identified an optimal three-cluster solution (K=3), which yielded the highest ARI across all analyses (0.179). However, this ARI value indicates poor agreement with clinical severity classifications, being substantially below the threshold for meaningful correspondence (ARI approaching 1.0). The silhouette score of 0.238 was well below the standard for good clustering quality (>0.5), indicating weak cluster cohesion and separation. Despite the poor quantitative metrics, visual inspection of the clustering heatmap revealed some discernible patterns: the upper two clusters (clusters 2 and 3) were predominantly composed of DHF patients (dark blue), while the bottom cluster (cluster 1) contained primarily healthy controls (white) and DF patients (light blue) (Figure 3B).

### Analysis of expression of ICOS and CD27 on T cells

Clustering analysis of ICOS+ combinations using hierarchical clustering with Ward’s linkage yielded a two-cluster solution (K=2) with an ARI of 0.166. The silhouette score of 0.138, again indicated weak clustering structure, far below acceptable thresholds for meaningful cluster quality. Although the clustering metrics were poor, visual inspection of the clustering heatmap (Figure 3C) revealed that cluster 1 appeared enriched for DHF patients, while cluster 2 contained mostly a mix of healthy controls and DF patients. However, the low ARI suggests that this pattern may not reflect genuine biological clustering corresponding to disease severity.

### Analysis of the expression of CLA on T cells

Clustering of the six markers of CLA+ combinations using hierarchical clustering with complete linkage resulted in a three-cluster solution (K=3) with the lowest ARI of 0.162, indicating minimal agreement with clinical classifications. The corresponding silhouette score was 0.214, which remained well below the threshold generally considered indicative of good clustering quality (>0.5). The clustering heatmap revealed that cluster 2 was primarily composed of healthy patients, cluster 3 was enriched for DHF patients, and cluster 1 mostly included a combination of DF and DHF patients (Figure 3D).

In summary, the unsupervised clustering analyses revealed poor correspondence between functionally defined T-cell subsets and clinical severity classifications. All ARI values (0.162-0.179) were substantially below meaningful thresholds indicating that the natural clustering patterns in the immunological data do not align with the predefined clinical groupings. Similarly, silhouette scores across all analyses (0.138-0.238) fell below the standard for good clustering quality (>0.5), suggesting weak internal cluster structure. Across all marker sets, hierarchical clustering yielded higher ARI values than k-means, and was therefore favored for interpretation, despite the overall low performance.

### Phenotypic differences in CD4+ and CD8+ T cells in lean and obese patients with acute dengue

As we observed differences in ex vivo IFNγ ELISpot responses in lean and obese patients (waist circumference ≥80 cm in women or ≥90 cm in men), we also sought to compare the differences in T cell phenotypes. We found that lean individuals had a significantly higher proportion of CD8+ T cells (p=0.04), while they had a significantly less proportion of CD4+ T cells (p=0.03) (Figure 4A). There were no differences in central memory (CCR7+CD62L+) CD4+ and CD8+ T cells or effector memory (CCR7-CD62L-) CD4+ and CD8+ T cells in lean or obese patients (Figure 4B). Although CD38 expression was higher in CD8+ T cells and specifically CD8+45RA− T cells, this was not significant (Figure 4C). CLA expression was highest in the CD8+CD45RA− of obese individuals, which was significantly higher compared to lean individuals (p=0.01), Figure 4D. While lean individuals had higher expression of CD27 on CD4+ T cells and lower expression of CD27 on CD8+ T cells this was not significant (Figure 4E). There were no differences in ICOS or CD127 expression of CD4+ or CD8+ T cells in obese vs lean patients (Figure 4E).

**Figure 4:**
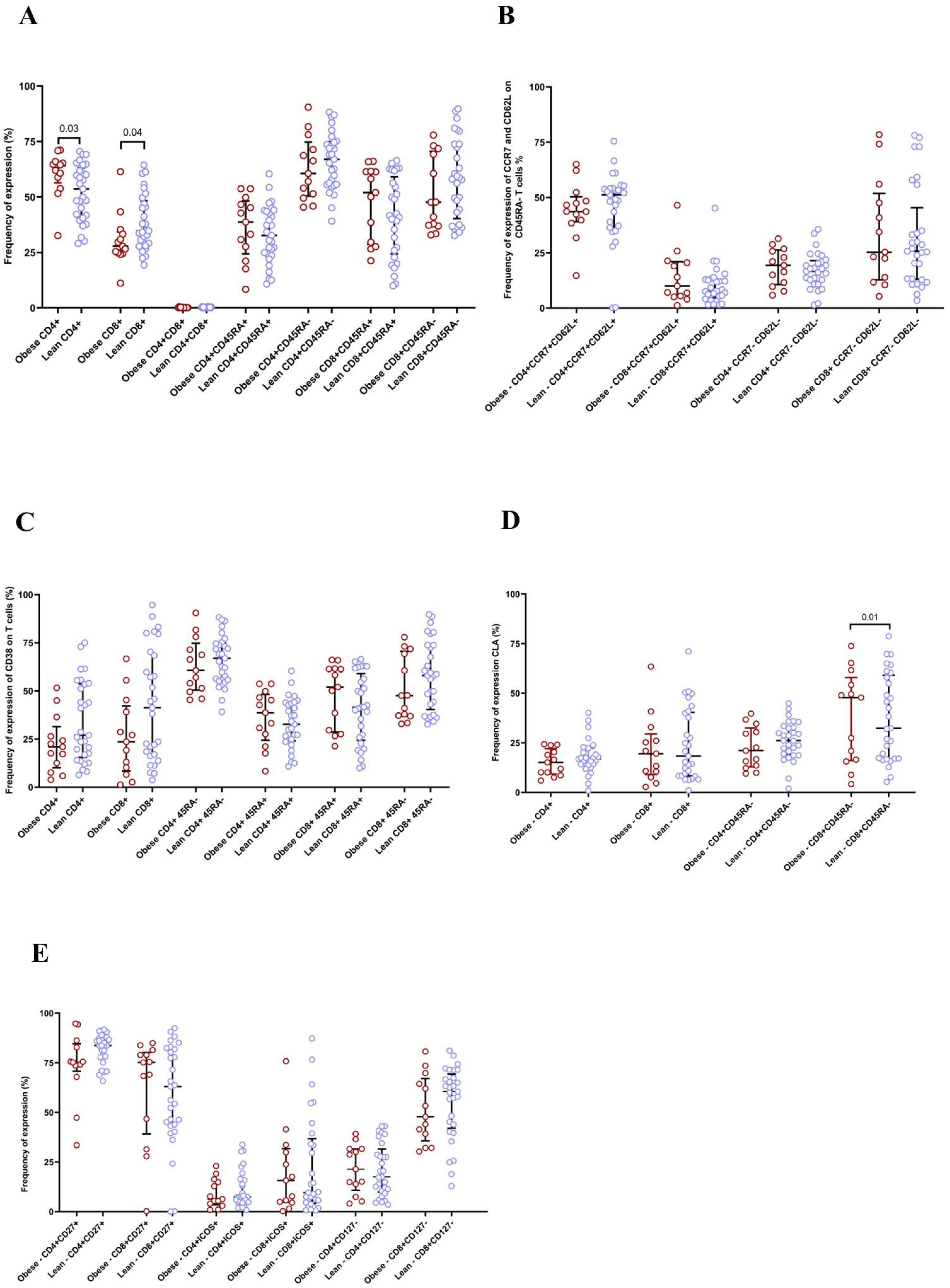
Differences in the phenotype of CD4+ and CD8+ T cells in obese and lean patients with acute dengue. CD4+ and CD8+ T cell subsets were phenotyped in patients with obese (waist circumferences (≥80 cm in women or ≥90 cm in men. Obese (n= 13 and lean (n= 29) patients recruited between days 6–8 from symptom onset by multicolour flowcytometry. The differences between the proportion of CD4+ CD8+ and CD45RA expressing T cells (A), expression of CCR7 and CD62L on CD45RA− T cells (B), CD38 expression (C), CLA expression (D) and CD27, CD127 and ICOS expression were assessed (E). The Mann Whitney U test (two-tailed) was used to calculate the differences between cohorts. The error bars indicate the median and the interquartile ranges.

### Phenotypic differences in CD4+ and CD8+ T cells patients based on different BMI with acute dengue

We also analyzed our data based on the BMI of patients, and all patients (BMI ≤23.9 and BMI >23.9). Individuals with a lower BMI (≤23.9), had a significantly higher proportion of central memory T cells (p=0.03) (Figure 5A). There was no difference in expression of CD38 on CD4+ and CD8+ T cells in patients with lower and higher BMI, although the CD38 expression was higher on CD8+CD45RA− T cells in patients with lower BMI, which was not significant (Figure 5B). The CLA expression was higher in patients with a higher BMI, in CD8+CD45RA− T cells, although this was not significant (Figure 5C). CD27 expression was significantly higher (p=0.03) on CD4+ T cells in patients with lower BMI (Figure 5D), while there was no difference in expression of ICOS and CD127.

**Figure 5:**
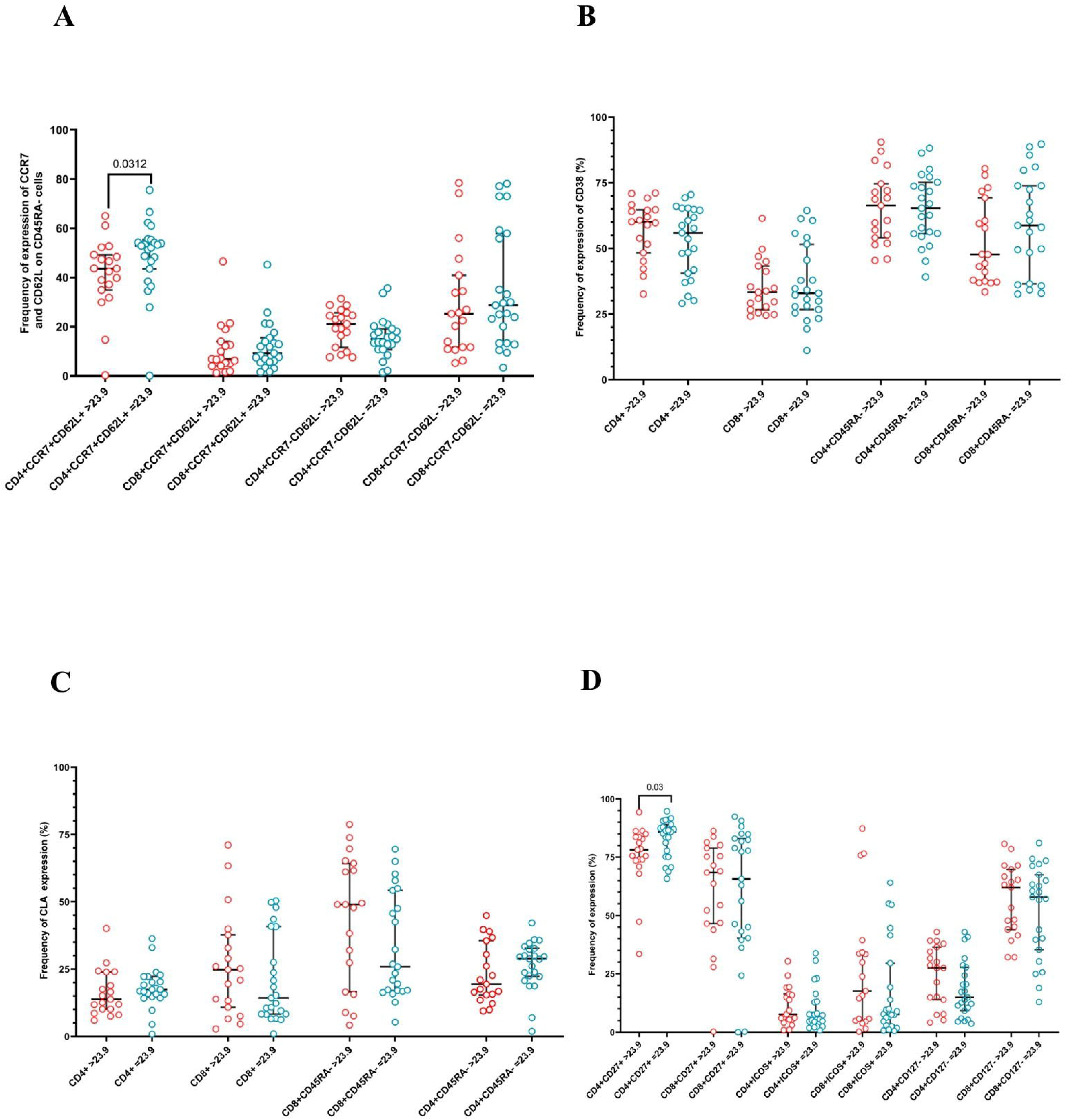
Differences in the phenotype of CD4+ and CD8+ T cells in patients with high (>23.9) and normal BMI (**≤**23.9) with acute dengue. CD4+ and CD8+ T cell subsets were phenotyped in patients with high BMI (n= 19) and normal BMI (n= 23) recruited between days 6–8 from symptom onset by multicolour flowcytometry. The differences between the proportion of CD4+ CD8+ and CD45RA expressing T cells (A), CD38 expression (B), CLA expression (C) and CD27, CD127 and ICOS expression were assessed (D). The Mann Whitney U test (two-tailed) was used to calculate the differences between cohorts. The error bars indicate the median and the interquartile ranges. The Mann– Whitney U test (two-tailed) was used to calculate the differences between cohorts. The error bars indicate the median and the interquartile ranges.

### Relationship between adiponectin levels, T cell activation and expression of co-stimulatory markers on CD4+ and CD8+ T cells

Similar to our previous study, we did not find any differences in adiponectin levels in patients with DF vs DHF (p= 0.97), or obese or lean patients (p= 0.25) [20]. However, as adiponectin has shown to regulate T cell proliferation and cytokine production and has many immunomodulatory actions on dendritic cells, NK cells and B cells [22,42], we sought to find out the associations of adiponectin levels with phenotypic differences in T cells. Although the patients enrolled in cohort 1 and 2 were different, in cohort 1 adiponectin was measured in time point F (≤ 4 days since onset of illness) and in cohort 2, between day 6 to 8. We found that adiponectin levels were significantly higher in in patients with DF in cohort 2 (p=0.009), compared to cohort 1, although no differences were seen in patients with DHF (Figure 6A). Interestingly, although adiponectin levels during early illness correlated with the frequency of DENV-specific ex vivo IFNγ producing CD4+ (Spearman’s r=0.37, p=0.02) and CD8+ (Spearman’s r=0.45, p=0.004), T cells at time point S (days 5 to 7 since onset of illness) in patients with DF, the adiponectin levels inversely correlated with IFNγ producing CD4+ (Spearman’s r=−0.74, p=0.04) and CD8+ (Spearman’s r=−0.86, p=0.01) in patients with DHF (Figure 6B).

**Figure 6:**
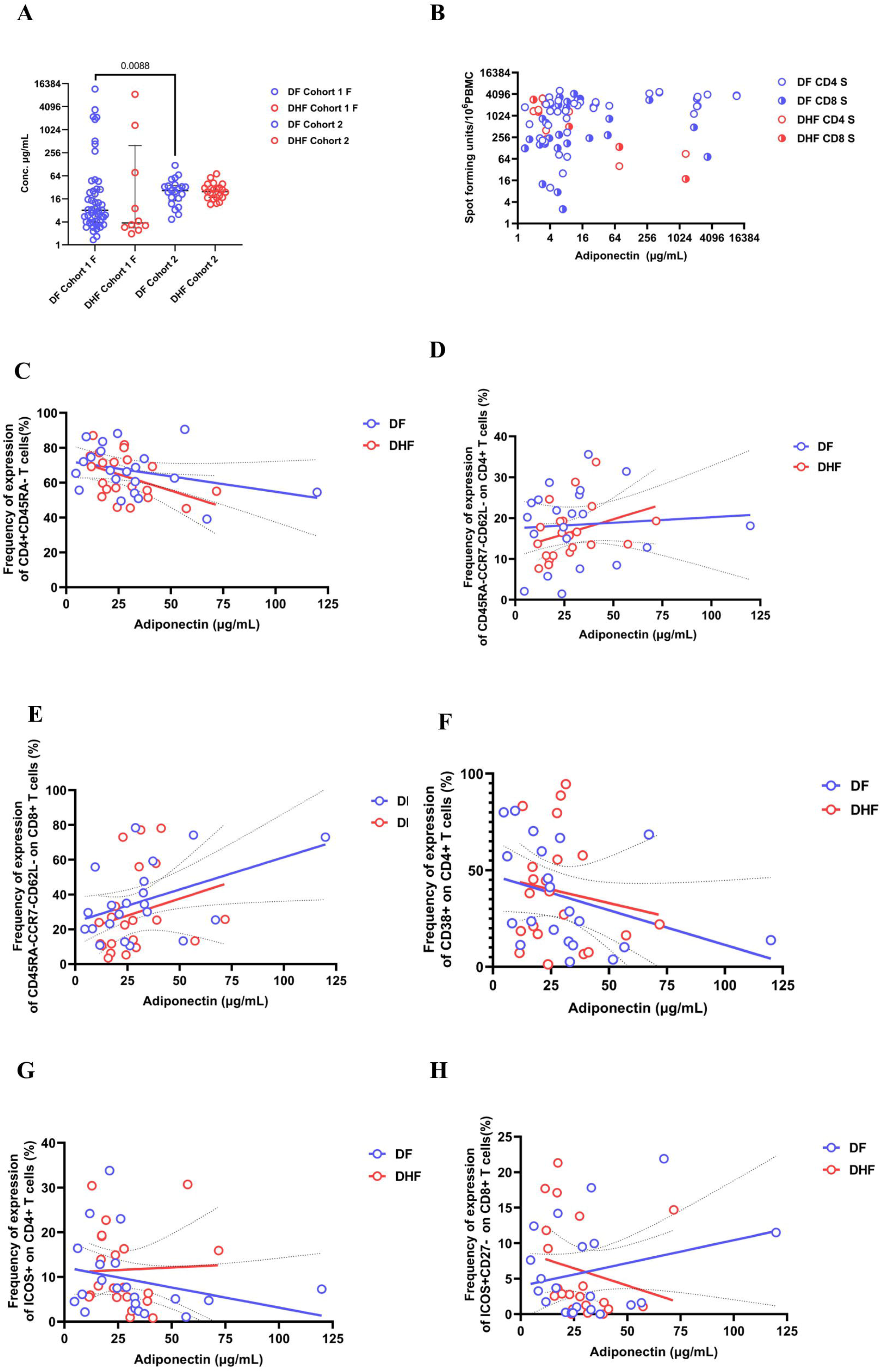
Associations between adiponectin levels and functional or phenotypic characteristics of CD4□ and CD8□ T cells in patients with dengue. Adiponectin levels were assessed in serum samples collected during the febrile phase (F: ≤4 days since symptom onset) in cohort 1 (n= 60) and between day 6 to 8 of illness in cohort 2 (n= 42). Adiponectin levels were compared between patients with DF and DHF between cohort 1 (time point F) and cohort 2 (A). In cohort 1, early adiponectin levels (F sample, ≤4 days since onset) were correlated with the frequency of DENV-specific IFNγ-producing CD4+ and CD8+ T cells at the second time point of illness (B). The adiponectin levels were correlated with CD4+CD45RA− T cells (C), CD4+CD45RA-CCR7-CD62L− T cells (D), CD8+CD45RA-CCR7-CD62L− T cells (E), CD4+CD38+ T cells (F), CD4+ICOS+ T cells (G) and CD8+ICOS+CD127− T cells (H). Group comparisons were performed using the two tailed Mann Whitney U test. Correlations were assessed by Spearman’s rank order correlation. Each point represents an individual patient and trend lines indicate linear regression with 95% confidence intervals.

While adiponectin levels also inversely correlated with CD4+CD45RA− cells in patients with DHF (Spearman’s r=−0.49, p=0.02) (Figure 6C), they correlated with CD4+ (Spearman’s r=0.46, p=0.03) and CD8+ (Spearman’s r=0.47, p=0.03) effector memory T cells (CD45RA-CCR7-CD62L−) (Figure 6D and 6E). No such correlation was seen in patients with DF. Instead, adiponectin levels inversely correlated with CD4+CD38+ T cells (Spearman’s r=−0.49, p=0.02), CD4+ICOS+ T cells (Spearman’s r=−0.44, p=0.04) in patients with DF (Figure 6F, G). In contrast, in patients with DHF, serum adiponectin levels were shown to negatively associate with CD8+ICOS+CD27− T cells (Spearman’s r=−0.52, p=0.01) (Figure 6H). There was no association between adiponectin with CLA or any other markers in patients with DF or DHF.

## Discussion

In this study we have assessed the functionality and phenotype of patients with DF and DHF and also compared responses in obese (waist circumference ≥80 cm in women or ≥90 cm in men) and lean patients with acute dengue, and also patients with high (>23.9) and normal BMI. We found that while DENV-specific IFNγ producing CD4+ and CD8+ T cells expanded from the first time point (≤4 days since onset of illness) to the second time point (5 to 7 days since onset of illness), in patients with DF, no such increase was seen in patients with DHF. Although we had investigated DENV specific T cell responses in patients with varying severity of dengue in our earlier studies, we and others had only included overlapping peptides representing DENV NS3, NS5 and NS1 of only one DENV serotype [13,40]. Here we used a pool of CD4+ and CD8+ T cell epitopes representing all DENV proteins and identified from all DENV serotypes. Although we did see patients with DF have a robust DENV specific T cell response evolve during the course of illness as in our previous study[40], we also observed that patients with DHF had higher frequencies of DENV-specific IFNγ producing CD8+ T cells as observed by Gregorova et al [13]. Therefore, patients who progress to develop DHF, appear to have a higher frequency of DENV-specific CD8+ T cells, which are possibly highly cross reactive in nature, but are possibly unable to sufficiently expand the IFNγ producing DENV-specific T cells, for the current infection required to eliminate the virus.

We saw that lean individuals also showed a significant increase in the frequency of both CD4+ and CD8+ DENV-specific T cell responses over time, with no such increase in obese individuals, suggesting that obesity may impair the functionality of DENV-specific T cell responses. These differences were seen when comparing individuals with central obesity and lean individuals, and differences were not seen when comparing T cell responses in individuals with high and normal BMI, possibly as we have shown that BMI is a poor indicator of the presence of visceral fat and underlying chronic inflammation [14]. Adiponectin is one of the adipokines which regulates T cell proliferation and cytokine production [22,42], are obese individuals typically have lower levels of adiponectin [15]. Adiponectin levels increased over time in patients with DF and was found to correlate with the increase in the frequency of DENV-specific IFNγ producing CD4+ and CD8+ T cell responses, whereas this was not seen in patients with DHF. Therefore, since we recently showed that obesity was associated with increased disease severity with higher levels of CRP, ferritin and liver transaminases, it would be important to further explore how obesity affects the functionality of T cell responses and disease severity, as polyfunctional DENV-specific T cell responses have shown to associate with milder illness [13,41].

Patients with acute dengue had an altered T cell phenotype with an increase in activated memory CD8+ T cells expressing skin homing markers compared to healthy individuals. The effector memory CD8+ T cells were significantly contracted in patients with DHF, with significant increases in CLA and ICOS with an increase in CD8+ T cells which had downregulated CD27 and CD127. We did not observe any differences in T cell phenotypes between lean and obese individuals, except higher frequency of CLA expressing CD8+CD45RA− memory T cells in obese individuals. In a previous study, although most DENV NS3 specific T cell restricted through HLA-A*1101 were shown to express CLA in patients with acute dengue, CLA expression was not shown to associate with disease severity and CLA expression was shown to be similar to non-dengue controls [29]. However, in our study we saw higher expression of CLA in patients with DHF, while CD8+ memory T cells of both patients with DF and DHF had significantly higher levels of CLA expression compared to healthy individuals. The discrepancy in results could be due to the patient numbers in the previous study (n=6) [29], compared to ours (n=42). In all our patients the CLA expressing CD8+ T cells were memory T cells and in patients with DHF almost all expressed CD38, suggesting that cross reactive CD8+ T cells specific to the previous DENV infection could be activated and homing to the skin. Although it is not clear if these CD8+CD45RA-CD38+CLA+ T cells are protective or cause pathogenesis (although significantly also high in obese individuals), it indicates that the skin is possibly an important site of viral replication, as many types of cells such as keratinocytes, dendritic cells and innate like lymphoid cells that reside in the skin have shown to be infected with the virus [9]. Indeed scRNA-seq studies have shown that both CD4+ and CD8+ T cells in patients with acute dengue upregulate genes encoding E-selectin and CLA during early illness, which further supports that the T cells home to the site of initial infection with the DENV [4].

ICOS expression was significantly upregulated in both CD4+ and CD8+ T cells in patients with DF and DHF, although the highest expression was seen in CD8+ T cells in patients with DHF. ICOS is upregulated upon T cell activation and regulates T cell proliferation and survival. While ICOS expression of CD4+ T cells are important for differentiation into follicular helper T cells, survival of CD4+ regulatory T cells and maintenance of effector and central memory CD4+ T cells, upregulation of ICOS expression on CD8+ T cells has shown to induced homing of CD8+ T cells in to non-lymphoid tissues [28]. The high expression of ICOS on CD8+ T cells especially in patients with DHF, further supports that the CD8+ T cells are likely to home to tissues (including the skin), where the primary site of virus replication occurs. However, we also found that CD27 and CD127 expressions were downregulated on CD4+ T cells in patients with DHF. CD27 is an important co-stimulatory molecule, which plays an important role in inducing antiviral responses and development of T cell memory [12]. CD127 downregulation has mainly been studies in the context of HIV infection, which associated with immune dysfunction and loss of CD4+ memory T cells [17]. Downregulation of both CD27 and CD127, especially in patients with DHF, may further contribute to impaired antiviral responses and development of virus specific memory T cell responses. Although comparison of different phenotypic markers of patients with DF and DHF provides insight into potential mechanisms of disease pathogenesis, they did not form distinct immunological signatures that correspond to dengue disease severity as currently classified, with the unsupervised PCA analysis. The results suggest either substantial immunological heterogeneity within clinical severity categories, or that additional unmeasured factors may be required to capture the immune determinants of dengue pathogenesis and severity.

Although we did not find many phenotypic differences in CD4+ and CD8+ T cells in obese vs lean individuals (except expression of CLA), serum adiponectin levels were found to inversely correlate with activation of CD8+ T cells and expression of ICOS by CD4+ T cells, while inversely correlating with ICOS expression and CD27 downregulation by CD8+ T cells. Therefore, adiponectin does seem to have varied effects on functionality and phenotypic characteristics of both CD4+ and CD8+ T cells in patients with DF and DHF, which should be further investigated to better understand the relevance.

In summary, in this study we have found that DENV-specific IFNγ producing CD4+ and CD8+ expanded during the course of illness in patients with DF, but not with DHF. Furthermore, while lean individuals also had a significant increase in the frequency of T cell responses during the course of the illness, this was not seen in obese patients. Patients with acute dengue had an altered T cell phenotype with an increase in activated memory CD8+ T cells expressing skin homing markers, which were also significantly elevated in obese patients. The effector memory CD8+ T cells were significantly contracted in patients with DHF, with a significant increase in ICOS expressing CD8+ T cells and CD27 and CD127 downregulation in CD4+ T cells, suggesting impaired functionality and phenotypic characteristic, especially in obese patients could contribute to disease pathogenesis.

## Data Availability

All data is available within the manuscript and supplementary files.

## Acknowledgements

We are grateful to the NIH, USA (grant number 5U01AI151788-02) and the UK Medical Research Council.

## Supplementary figure legends

**Supplementary Figure 1 (S1): Gating strategy used for identifying CD4+ and CD8+ subsets and determining expression of CD45, CCR7, CD62L, CD38, and CLA**

CD45+ leukocytes were initially gated followed by the exclusion of cell doublets using FSC-A vs FSC-H and SSC-A vs SSC-H. Viable cells were then selected and monocytes (CD14+), B cells (CD19+) and granulocytes (CD66b+) were removed using a dump channel. The remaining CD3+ T cells were subsequently divided into CD4+ and CD8+ subsets based on their respective surface marker expression. Within each subset cells were assessed for CD38 as an activation marker and CLA, a marker associated with skin-homing potential. T cells were then divided into naive and memory populations based on CD45RA expression and each group was further analyzed for CD38 and CLA expression. To define memory subsets more precisely CD45RA+ and CD45RA− populations were subdivided using CCR7 and CD62L expression to identify naive T cells (TN), central memory (TCM), effector memory (TEM), and terminally differentiated effector memory cells (TEMRA) within both CD4+ and CD8+ T cell compartments.

**Supplementary Figure 2 (S2): Gating strategy used for identifying CD4+ and CD8+ subsets and determining expression of CD27, CD127 and ICOS**

Leukocytes were first gated based on CD45 expression. Then doublets were excluded using FSC-A vs FSC-H and SSC-A vs SSC-H. Live cells were then selected and non-T cells including monocytes (CD14+) B cells (CD19+) and granulocytes (CD66b+) were removed using a dump channel. CD3+ T cells were identified and further divided into CD4+ and CD8+ subsets. Within these subsets the expression of CD27 and CD127 was examined individually and their co expression was used to define distinct T cell differentiation stages. ICOS expression was also analyzed to identify activated T cells with co-stimulatory potential.

**Supplementary Figure 3 (S3): Frequency of expression of CD32 and CD64 on CD4+ and CD8+T cells in in patients with varying severity of acute dengue**

The frequency of CD32+ and CD64+ in CD4+ and CD8+ T cell populations was assessed in patients with dengue fever (DF), dengue hemorrhagic fever (DHF), and healthy controls (HC). Across all groups CD32 and CD64 expression was either very low or undetectable in both CD4+ and CD8+ T cells. There were no significant differences in the frequency of CD32+, CD64+ or CD32+CD64+ co expressing cells among DF, DHF or HC groups suggesting that these receptors are not prominently expressed on T cells in the context of acute dengue. The Mann Whitney U test (two-tailed) was used to assess statistical differences between groups. Data are presented as individual values with bars indicating the median and interquartile ranges.

