## Supplementary methods for "Functionality and phenotype of T cells in patients with varying severity of acute dengue and metabolic status"

**Data Preprocessing and Standardization**

Flow cytometry data comprising immune marker expression profiles were analyzed using Python version 3.11.5. Samples containing missing values were excluded via complete case analysis. All continuous variables were standardized using z-score normalization to ensure uniform scaling across features for multivariate analysis. Disease severity was used as the primary grouping variable in downstream comparisons.

**Multicollinearity Assessment**

Multicollinearity was assessed using pairwise Pearson correlation coefficients and variance inflation factor (VIF) scores. Markers with VIF > 10 or pairwise |r| > 0.8 were considered collinear and evaluated for exclusion. Final marker retention was guided by statistical and biological criteria to minimize redundancy while preserving dimensionality.

The following flow cytometry markers were excluded:
CD4+CD45RA+, CD4+CD38−, CD4+CD45RA+CD38+, CD4+CD45RA+CLA+, CD4+CD27+CD127+, CD4+CD32+, CD4+CD64+, CD4+CD32+CD64+, CD8+CD45RA+, CD8+CD38−, CD8+CD45RA+CD38+, CD8+CD45RA+CLA+, CD8+CD27+CD127+, CD8+CD32+,CD8+CD64+,CD8+CD32+CD64+.
Following exclusion of samples with missing data and removal of collinear markers while considering biologically important ones, the final dataset comprised 51 samples and 33 immune markers. This reduced dataset was used for all downstream analyses, including PCA.

**Principal Component Analysis**

An exploratory principal component analysis (PCA) was conducted using the scikit-learn library in Python on z-score standardized flow cytometry marker data. The primary objective was to evaluate whether samples grouped by disease severity, exhibited separation in reduced-dimensional space. Variance explained by each principal component was computed, and a scree plot was used to guide the selection of components for visualization. PC1 and PC2, which explained 26.31% and 12.55% of the variance respectively, were selected for 2D visualization. Principal component scores were plotted in a scatter plot with samples color-coded by disease severity. Axis labels indicated the proportion of variance explained by each component.

**Clustering Analysis**

Unsupervised clustering analysis was conducted to identify distinct immune phenotypes based on standardized flow cytometry marker data. All clustering analyses and visualizations were conducted in R (version 4.3.2) using the cluster, factoextra, pheatmap, and ggplot2 packages. Clustering was performed separately for three biologically relevant T-cell marker subsets, each representing functionally distinct immune states involved in dengue pathogenesis:

Skin-homing memory T cells (6 markers): *CD4+CD45RA-CLA+, CD8+CD45RA-CLA+, CD4+CLA+, CD8+CLA+, CD4+CD38+CLA+, CD8+CD38+CLA+*

Exhaustion markers (6 markers): *CD4+CD127-, CD8+CD127-, CD4+CD27-, CD8+CD27-, CD4+CD27-CD127-, CD8+CD27-CD127-*

Activation-associated markers (8 markers): *CD4+ICOS+, CD8+ICOS+, CD4+ICOS+CD27-, CD4+ICOS+CD127-, CD8+ICOS+CD27-, CD8+ICOS+CD127-, CD4+ICOS+CD27-CD127-, CD8+ICOS+CD27-CD127-*

Two complementary unsupervised clustering approaches were applied for each marker subset:

K-means clustering was performed using the kmeans() function in R, with nstart = 25 and set.seed(123) to ensure reproducibility. The number of clusters (K) was selected based on a combination of the Elbow method (evaluating within-cluster sum of squares) and Silhouette scores (assessing average silhouette width). K values of 2, 3, and 4 were evaluated.

Hierarchical clustering was conducted using Euclidean distance and three linkage methods: Ward’s method (ward.D2), complete linkage, and average linkage. Dendrograms were generated to visualize cluster structure and guide cluster cut-point selection.

Cluster validation was performed using the following metrics:

Adjusted Rand Index (ARI): The primary metric, evaluating agreement between derived clusters and predefined clinical groups (e.g., severity categories), adjusted for chance.

Silhouette Score: A secondary metric measuring cluster cohesion and separation (range: –1 to 1), used to support interpretability.

The optimal clustering configuration for each analysis was primarily determined based on the highest ARI. In cases where ARI values were comparable across methods, silhouette scores were additionally considered to support interpretability.
