## Supplementary figure for "Functionality and phenotype of T cells in patients with varying severity of acute dengue and metabolic status"

**Supplementary Figure 1 (S1) – Panel 1**

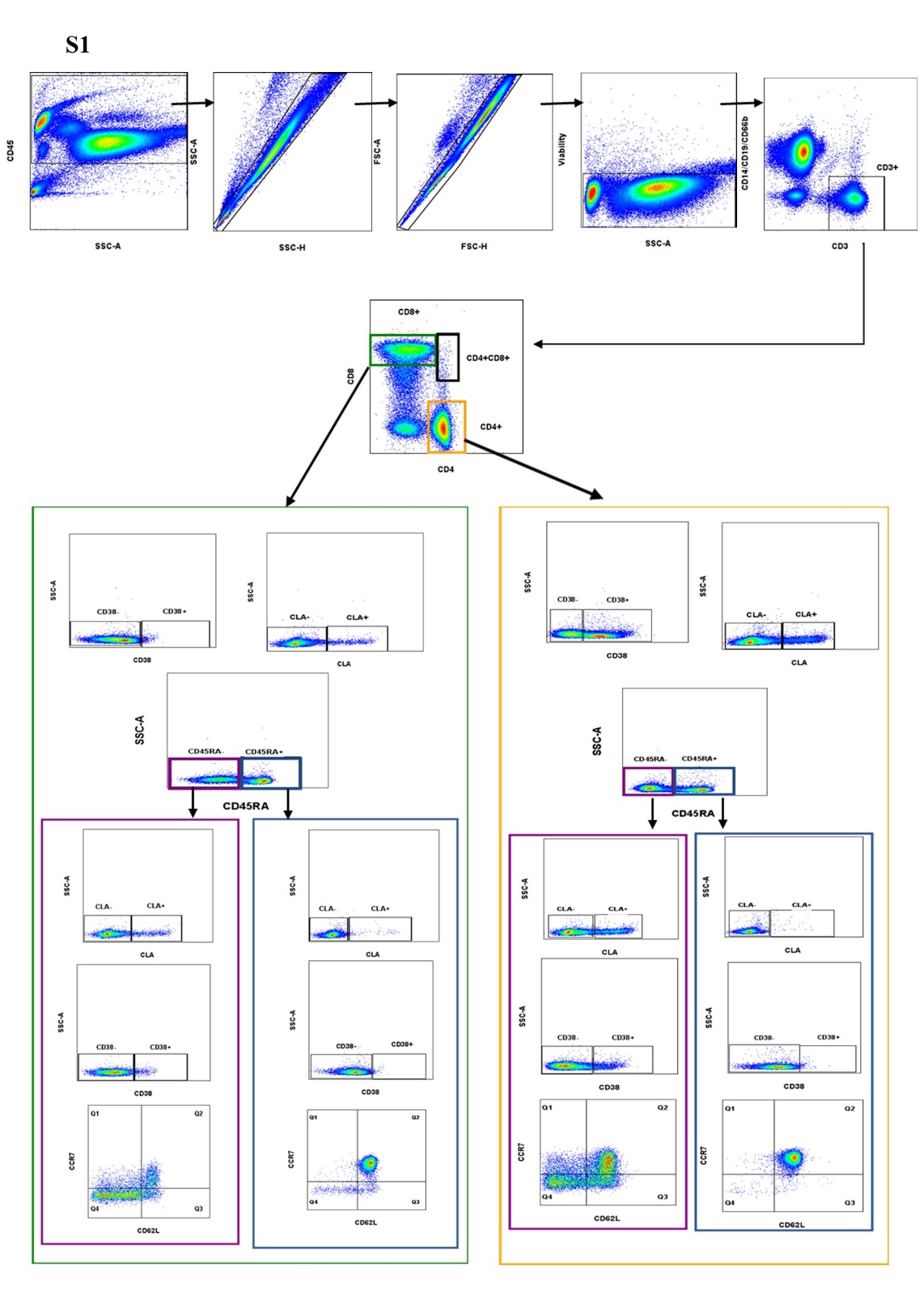

**S1 – Figure legend**

CD45+ leukocytes were initially gated followed by the exclusion of cell doublets using FSC-A vs FSC-H and SSC-A vs SSC-H. Viable cells were then selected and monocytes (CD14+), B cells (CD19+) and granulocytes (CD66b+) were removed using a dump channel. The remaining CD3+ T cells were subsequently divided into CD4+ and CD8+ subsets based on their respective surface marker expression. Within each subset cells were assessed for CD38 as an activation marker and CLA, a marker associated with skin-homing potential. T cells were then divided into naive and memory populations based on CD45RA expression and each group was further analyzed for CD38 and CLA expression. To define memory subsets more precisely CD45RA+ and CD45RA- populations were subdivided using CCR7 and CD62L expression to identify naive T cells (TN), central memory (TCM), effector memory (TEM), and terminally differentiated effector memory cells (TEMRA) within both CD4+ and CD8+ T cell compartments.

**Supplementary Figure 2 (S2) – Panel 2**

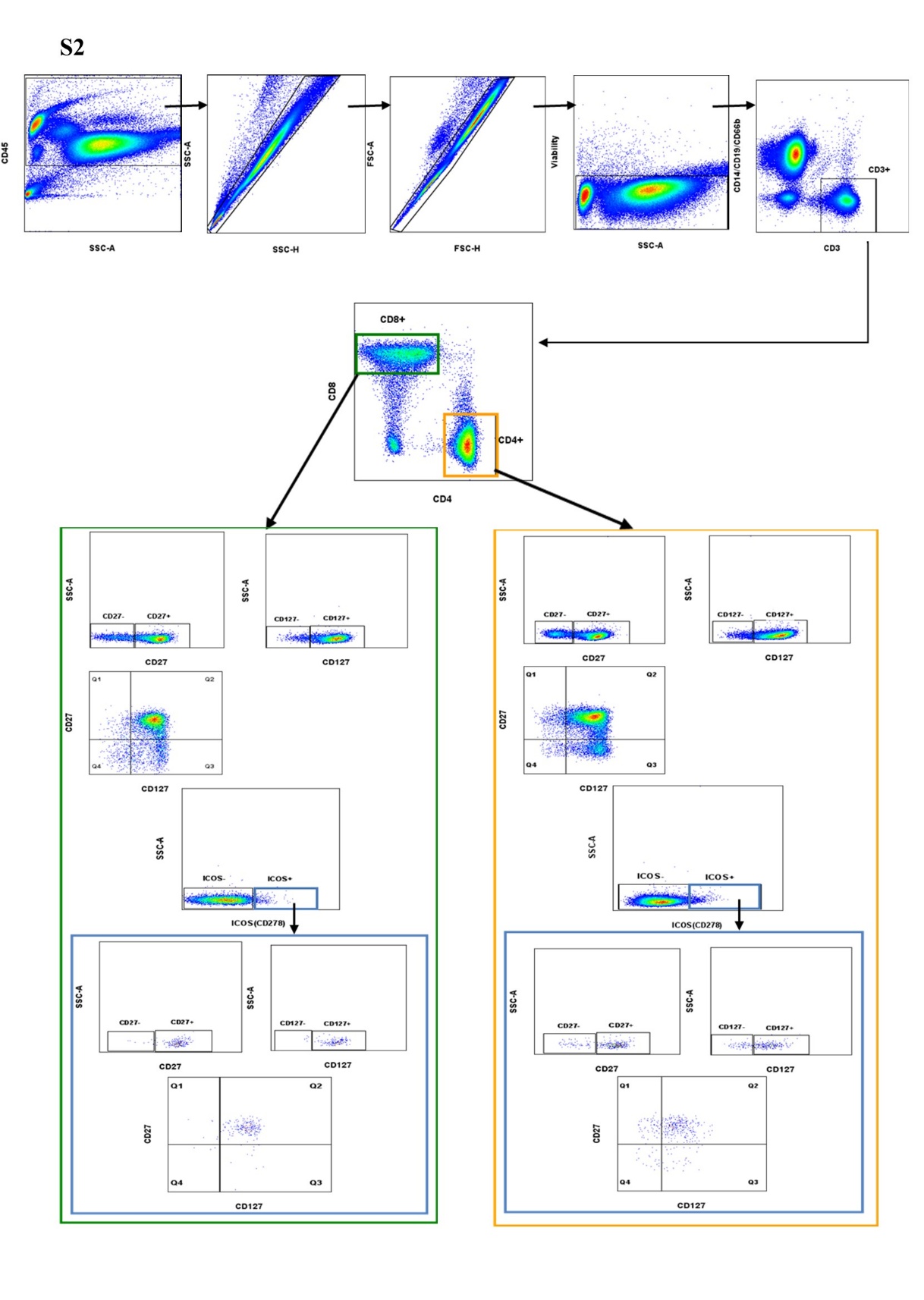

**S2 – Figure legend**

Leukocytes were first gated based on CD45 expression. Then doublets were excluded using FSC-A vs FSC-H and SSC-A vs SSC-H. Live cells were then selected and non-T cells including monocytes (CD14+) B cells (CD19+) and granulocytes (CD66b+) were removed using a dump channel. CD3+ T cells were identified and further divided into CD4+ and CD8+ subsets. Within these subsets the expression of CD27 and CD127 was examined individually and their co expression was used to define distinct T cell differentiation stages. ICOS expression was also analyzed to identify activated T cells with co-stimulatory potential.

**Supplementary Figure 3 (S3) – Frequency of expression of CD32 and CD64 on CD4+ and CD8+T cells**

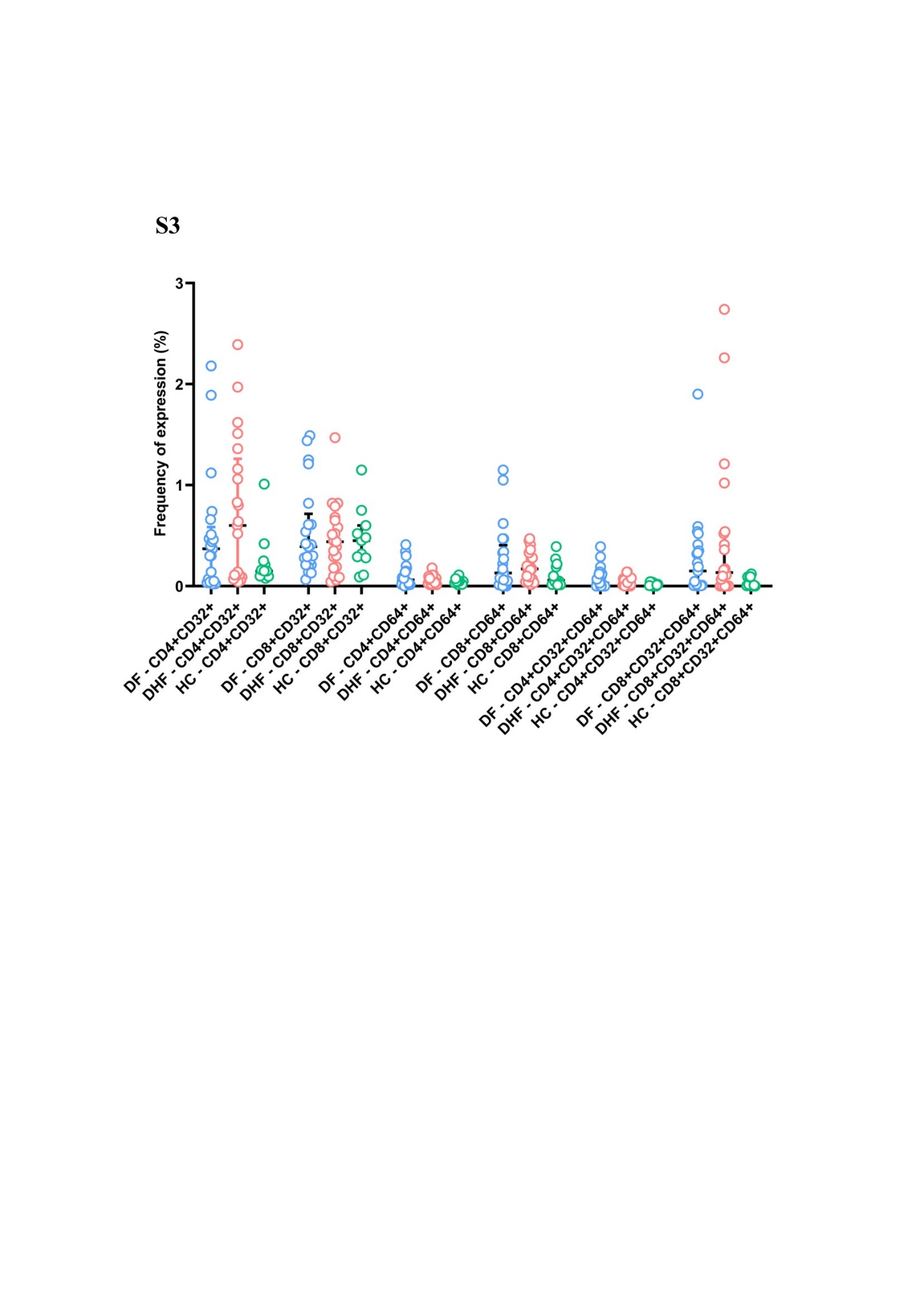

**S3 – Figure legend**

The frequency of CD32+ and CD64+ in CD4+ and CD8+ T cell populations was assessed in patients with dengue fever (DF), dengue hemorrhagic fever (DHF), and healthy controls (HC). Across all groups CD32 and CD64 expression was either very low or undetectable in both CD4+ and CD8+ T cells. There were no significant differences in the frequency of CD32+, CD64+ or CD32+CD64+ co expressing cells among DF, DHF or HC groups suggesting that these receptors are not prominently expressed on T cells in the context of acute dengue. The Mann Whitney U test (two-tailed) was used to assess statistical differences between groups. Data are presented as individual values with bars indicating the median and interquartile ranges.
