## Supplementary data for "Functionality and phenotype of T cells in patients with varying severity of acute dengue and metabolic status"

|  | **DF**  **N = 50** | **DHF**  **N = 12** | **HC**  **N = 19** |
| --- | --- | --- | --- |
| **Gender** | | | |
| Female | 23 (46%) | 5 (41.6%) | 11 (57.8%) |
| Male | 27 (54%) | 7 (58.4%) | 8 (42.2%) |
| **Age** | | | |
| Mean (SD) | 30.8 (16.3) | 25.0 (6.9) | 32.3 (6.5) |
| **BMI** | | | |
| Mean (SD) | 23.1 (5.3) | 24.8 (4.9) | 23.8 (2.5) |
| **Waist circumference** | | | |
| Mean (SD) | 77.0 (23.7) | 71.33 (24.5) | 78.0 (11.2) |
| **PCV** | | | |
| Mean (SD) | 40.9 (3.4) | 42.12 (5.4) | N/A |
| **Platelets (x10^9^/L)** | | | |
| Mean (SD) | 83.3 (49.4) | 32.5 (21.19) | N/A |
| **AST** | | | |
| Mean (SD) | 107.2 (115.7) | 188.5 (159.4) | N/A |
| **ALT** | | | |
| Mean (SD) | 102.8 (123.2) | 139.1 (100.7) | N/A |
| **CRP** | | | |
| Mean (SD) | 42.5(55.5) | 35.3 (29.1) | N/A |
| **Days of fever** | | | |
| Mean (SD) | 3.6 (0.5) | 3.8 (0.3) | N/A |
| **Serotype** | | | |
| DEN1 | N/A | N/A | N/A |
| DEN2 | 7 | 1 | N/A |
| DEN3 | 6 | 4 | N/A |
| **NS1 rapid test +** | 32 | 8 | N/A |

**Supplementary data**

**Table 1** Demographic data and clinical characteristics of the study cohorts 1(Functional)

**Table 2** Demographic data and clinical characteristics of the study cohort 2 (Phenotypic)

|  | **DF**  **N = 21** | **DHF**  **N = 21** | **HC**  **N = 11** |
| --- | --- | --- | --- |
| **Gender** | | | |
| Female | 12(57.1%) | 8(38%) | 6(54.5%) |
| Male | 9(42.9%) | 13(62%) | 5(45.5%) |
| **Age** | | | |
| Mean (SD) | 39.6(13.4) | 39.1(15.8) | 30.2(5.3) |
| **BMI** | | | |
| Mean (SD) | 24.8(5.9) | 25.5(5.8) | 22.8(3.9) |
| **Waist circumference** | | | |
| Mean (SD) | 83.4(9.5) | 83.4(10.4) | 79.5(12.2) |
| **PCV** | | | |
| Mean (SD) | 40.1(5.2) | 40.5(5.9) | N/A |
| **Platelets (x10^9^/L)** | | | |
| Mean (SD) | 100.4(64.0) | 30.7(32.7) | N/A |
| **AST** | | | |
| Mean (SD) | 123.1(152.3) | 290.8(431) | N/A |
| **ALT** | | | |
| Mean (SD) | 92.6(95.0) | 150.4(126.7) | N/A |
| **CRP** | | | |
| Mean (SD) | 16.1(18.3) | 29.8(23.9) | N/A |
| **Days of fever** | | | |
| Mean (SD) | 6.8 (0.8) | 6.5(0.9) | N/A |
| **NS1** | 2 | 7 | N/A |
| **Dengue IgG/IgM** | 21 | 21 | N/A |
